# Combination of heart failure and atrial fibrillation worsens ethnicity-related disparity: An individual patient-level meta-analysis of randomised trials

**DOI:** 10.1101/2025.07.31.25332510

**Authors:** Sebastian J Fox, Asgher Champsi, Evan J Hardy, Karina V Bunting, Giuseppe Rosano, Michael Böhm, Marcus D. Flather, Dipak Kotecha, the cardAIc group, the Beta-blockers in Heart Failure Collaborative Group

## Abstract

**Aims:** Ethnicity is known to influence patient outcomes and treatment efficacy, but knowledge is limited how multimorbidity interacts with clinical events, for example when heart failure (HF) and atrial fibrillation (AF) combine.

**Methods and Results:** 16,713 patients were included from 12 randomized placebo-controlled trials in HF (11 versus beta-blockers and 1 versus spironolactone), of which 13,568 patients (81.2%) were in sinus rhythm and 3,145 (18.8%) had comorbid AF at baseline. Non-white ethnicity was recorded in 1,899 (11%), with these patients being younger (median age 58 versus 67 years) and suffering higher rates of diabetes, hypertension, and lower left-ventricular ejection fraction (median 25% versus 30%) than those of white ethnicity. During median follow-up of 1.4 years (IQR 0.8-2.3), the primary outcome of all-cause mortality occurred in 394 (21%) non-white patients and 2,142 (15%) white patients, with confounder-adjusted hazard ratio (HR) 1.36, 95% CI 1.20-1.54; p<0.001. The impact of ethnicity on death was greater in patients with coexisting HF and AF (non-white versus white HR 2.05; 95% CI 1.55-2.70; p<0.001) than in those with HF in sinus rhythm (HR 1.24; 95% CI 1.08-1.41; p=0.002). The interaction p-value was 0.003, and confirmed using propensity-score matching to account for baseline differences (p=0.009). Similar disparities for the combination of ethnicity and comorbidity were seen for the secondary outcomes cardiovascular and HF-related death, and cardiovascular and HF-related hospitalisation.

**Conclusion:** Non-white patients with HF suffer from substantially higher rates of death, and comorbid AF leads to significant worsening of this ethnicity-related disparity.

## Introduction

Heart failure (HF) and atrial fibrillation (AF) are two emerging public health concerns due to rapid increases in prevalence and their association with substantial morbidity and mortality. The conditions frequently coexist^1^, and patients with concomitant HF and AF may not respond to conventional guideline-recommended therapy leading to even higher rates of adverse outcomes.^2, 3^ On a background of numerous patient-specific factors, ethnicity has long been considered an important contributor to variation in prognosis and treatment efficacy.^4^ The term ethnicity encompasses biological parameters and cultural attributes, with health perception, health behaviour and healthcare access all contributing to differences in clinical outcomes.^5^ These factors affect how patients present, the treatments they receive and their follow-up, making direct ethnicity comparisons challenging. Knowledge on the true impact of these issues on prognosis in patients with either HF or AF is available but limited.^6, 7^ In contrast, there is no consistent information on ethnicity in patients that have both HF and AF, with two observational studies finding conflicting results.^8, 9^

This study was designed to examine if the impact of ethnicity on clinical events is compounded by multimorbidity. The null hypothesis tested was of no difference in death (and other cardiovascular events) between non-white and white patients when comparing HF in sinus rhythm and HF with concomitant AF. Individual patient-level data from randomised controlled trials (RCTs) were utilised to reduce bias related to differences in definitions, healthcare delivery and outcome ascertainment that are commonly encountered in observational research.

## Methods

### Ethics

The study complies with the Declaration of Helsinki and the research proposal was approved by the University of Birmingham Ethical Review Committee (ERN_20-0647).

### Study selection and data harmonisation

Anonymised individual patient data were obtained from 12 RCTs comprising patients with HF, with or without concomitant AF (Supplementary Figure S1). All trials had appropriate ethical approvals, and a low risk of bias evaluated using the Cochrane Collaborations Risk of Bias Tool (Supplementary Figure S2). Trial data were obtained from the Beta Blockers in Heart Failure Collaborative Group (BB-meta-HF; Clinicaltrials.gov NCT0083244^10^) and the National Heart, Lung and Blood Institute Biologic Specimen and Data Repositories Information Coordinating Centre (BioLINCC).^11, 12^ A process of data harmonisation and data quality assessment was undertaken to combine all trial datasets using an established and robust approach.^2, 13^

### Population

The combined population from the 12 RCTs was divided into two groups according to cardiac rhythm status (sinus rhythm or AF) determined on the baseline electrocardiogram. Atrial flutter was not always indicted separately and so was included in the AF group, and those with a missing ECG or paced rhythm were excluded. Coding for each participant’s ethnicity was accepted from each corresponding trial database, and participants with missing ethnicity excluded. The categories of ethnicity were then collapsed into non-white and white groups. To avoid introducing bias for data not missing at random, no data imputation methods were used.

### Outcomes

The primary outcome for this study was all-cause mortality.

Secondary outcomes were cardiovascular (CV)-related mortality, HF-related mortality, CV-related hospitalisation and HF-related hospitalisation. Each trial performed independent adjudication of clinical outcomes.

### Statistical Analysis

Baseline data were summarised with mean and standard deviation (SD), median and interquartile range (IQR), or frequency and percentage. Differences between groups were analysed using a T-test, Mann-Whitney U test, or Chi-squared test. Incidence rates per 1000 person-days and incidence rate ratios were used to compare outcomes by ethnicity. Time-to-event analyses using Cox proportional hazard ratio (HR) models was the primary analysis, with proportionality testing used to confirm the hazards were proportional over time. Regression models included adjustment for age, gender, New York Heart Association (NYHA) class, left ventricular ejection fraction (LVEF), previous myocardial infarction (MI), diabetes and hypertension. Two interactions were found to be significant and included in the model: age and LVEF, and LVEF and NYHA class. To account for observed baseline differences between the ethnicity groups, a propensity-matched analysis was performed as a sensitivity analysis. The propensity-matched model employed one-to-one nearest neighbour matching with replacement.^14^ This generated an average treatment effect on the treated, summarising the difference in probabilities of an event occurrence between non-white patients and their propensity-matched white counterparts. Odds ratios were obtained by performing a logistic regression on the propensity-score matched groups. An exploratory analysis of treatment efficacy stratified by ethnicity was also performed, but only where randomised treatment effects were significant (within each rhythm group). This was done using an intention-to-treat approach according to the randomised allocation, regardless of treatment discontinuation or cross-over. All statistical analyses were performed using Stata (version 17, StataCorp LP, Texas) and a 2-tailed p-value of <0.05 was used to denote statistical significance.

## Results

The study included 12 RCTs of patients with HF (Supplementary Figure S1), including 10 trials with predominantly reduced LVEF (<40%),^12, 15-23^ 1 trial with a range of LVEF,^24^ and 1 trial with LVEF ≥45%.^11^ In total, individual patient data from 16,713 participants with HF were analysed, of which 13,568 (81.2%) were in sinus rhythm at baseline and 3,145 (18.8%) in AF. Non-white ethnicity was recorded in 1,899 (11%) and white in 14,817 (89%) (Supplementary Table S1 for breakdown of ethnicity by rhythm status). There were significant differences across baseline characteristics between non-white and white patients (Table 1; Supplementary Tables S2 and S3 for demographics by rhythm status). The non-white cohort were younger, with median age 58.0 years (IQR 49.0-67.0) versus 67.0 years for the white group (IQR 58.0-73.7; p<0.001). Non-white participants had a lower rate of prior MI, more hypertension and diabetes, lower median LVEF (25% [IQR 19%-34%]) versus 30% [IQR 22%-38%]), and were typically more symptomatic (NYHA class III/IV 72.1% versus 54.3%). There was no difference in the allocation to intervention or placebo with regards to ethnicity (p=0.31). For the propensity-matched analyses, the average treatment effect on the treated was estimated in 1,354 non-white patients and 1,011 white patients after baseline characteristics were balanced across groups (p>0.05 for all included covariates; Supplementary Table S4).

**Table 1:** Baseline demographics by ethnicity.

| Baseline characteristic | Non-white<br>(n=1,899) | White<br>(n=14,814) | p-value |
| --- | --- | --- | --- |
| Intervention arm, n (%) | 986 (51.9%) | 7510 (50.7%) | 0.31 |
| Heart rhythm |  |  |  |
| Sinus rhythm, n (%) | 1703 (89.7%) | 11865 (80.1%) | <0.001 |
| Atrial fibrillation, n (%) | 196 (10.3%) | 2949 (19.9%) | <0.001 |
| Female gender, n (%) | 634 (33.3%) | 4089 (27.6%) | <0.001 |
| Age, median years (IQR) | 58.0 (49.0-67.0) | 67.0 (58.0-73.7) | <0.001 |
| BMI, median kg/m<sup>2</sup> (IQR) | 28.0 (24.2-32.9) | 27.1 (24.4-30.6) | <0.001 |
| Systolic BP, median mmHg (IQR) | 120 (108-134) | 125 (112-140) | <0.001 |
| Diastolic BP, median mmHg (IQR) | 75 (68-82) | 77 (70-82) | <0.001 |
| Heart rate, mean (SD) | 82.0 (13.8) | 79.1 (12.8) | <0.001 |
| LVEF, median (IQR) | 0.25 (0.19-0.34) | 0.30 (0.22-0.38) | <0.001 |
| NYHA Class III/IV, n (%) | 1362 (72.1%) | 8039 (54.5%) | <0.001 |
| Comorbidities |  |  |  |
| Previous myocardial infarction, n (%) | 662 (34.9%) | 7905 (53.4%) | <0.001 |
| Previous stroke, n (%) | 73 (7.1%) | 693 (6.6%) | 0.56 |
| Previous CABG or PCI, n (%) | 342 (18.1%) | 3202 (22.2%) | <0.001 |
| Hypertension, n (%) | 1314 (70.5%) | 7697 (53.2%) | <0.001 |
| Peripheral arterial disease, n (%) | 156 (8.6%) | 1180 (8.3%) | 0.73 |
| Diabetes mellitus, n (%) | 747 (40.3%) | 3755 (25.9%) | <0.001 |
| Medications |  |  |  |
| Diuretic, n (%) | 1509 (93.7%) | 10387 (82.7%) | <0.001 |
| ACE inhibitor or ARB, n (%) | 1543 (95.8%) | 11874 (94.5%) | 0.036 |
| Aldosterone antagonist, n (%) | 67 (4.7%) | 1243 (8.6%) | <0.001 |
| Calcium channel blocker, n (%) | 202 (10.9%) | 1630 (11.2%) | 0.66 |
| Statin, n (%) | 451 (24.8%) | 3625 (25.6%) | 0.44 |
| Digoxin, n (%) | 1346 (72.2%) | 7221 (49.9%) | <0.001 |
| Oral anticoagulant, n (%) | 660 (35.4%) | 4592 (31.7%) | 0.002 |

IQR = interquartile range; BMI = body mass index; BP = blood pressure; SD = standard deviation; LVEF = left ventricular ejection fraction; NYHA = New York Heart Failure Association; CABG = coronary artery bypass grafting; PCI = percutaneous coronary intervention; ACE = angiotensin-converting enzyme; ARB = angiotensin receptor blocker. Refer to Supplementary Table S2, S3 for missing data.

### Primary outcome: All-cause mortality

During median follow-up of 1.4 years (IQR 0.8-2.3), death occurred in 394 (20.7%) non-white patients and 2,142 (14.5%) white patients. The adjusted HR of all-cause mortality for non-white versus white was 1.36, 95% CI 1.20-1.54; p<0.001. When stratified according to rhythm status, non-white HF patients with concomitant AF had the highest rate of all-cause mortality (Figure 1). The non-white versus white adjusted HR for all-cause mortality in patients with HF in sinus rhythm was 1.24 (95% CI 1.08-1.41; p=0.002) and 2.05 for those with HF plus AF (95% CI 1.55-2.70, p<0.001); Table 2. The differential impact of ethnicity was significantly greater with HF and AF combined (interaction p=0.003). Results were confirmed in the propensity score-matched analysis (p=0.12 for HF in sinus rhythm; p=0.001 for HF plus AF; p for comparison=0.009; Supplementary Table S5).

**Table 2:** Primary and secondary outcomes.

| Non-white vs white: | Crude event numbers (%) | Incidence rate ratio (95% CI), p-value | Adjusted hazard ratio (95% CI), p-value | Interaction p-value for additional comorbidity |
| --- | --- | --- | --- | --- |
| All-cause mortality |  |  |  |  |
| HF in sinus rhythm | 330/1703 (19.4%) vs 1610/11865 (13.6%) | 1.40 (1.23-1.58), p<0.001 | 1.24 (1.08-1.41), p=0.002 | 0.003 |
| HF plus AF | 64/196 (32.7%) vs 532/2949 (18.0%) | 1.85 (1.39-2.42), p<0.001 | 2.05 (1.55-2.70), p<0.001 |  |
| Cardiovascular-related mortality |  |  |  |  |
| HF in sinus rhythm | 278/1703 (16.3%) vs 1317/11865 (11.1%) | 1.42 (1.24-1.63), p<0.001 | 1.22 (1.05-1.41), p=0.007 | 0.14 |
| HF plus AF | 45/196 (23.0%) vs 432/2949 (14.6%) | 1.54 (1.09-2.21), p=0.012 | 1.63 (1.17-2.27), p=0.004 |  |
| Heart failure-related mortality |  |  |  |  |
| HF in sinus rhythm | 75/1703 (4.4%) vs 376/11865 (3.2%) | 1.38 (1.06-1.77), p=0.016 | 1.16 (0.89-1.52), p=0.28 | 0.24 |
| HF plus AF | 15/196 (7.7%) vs 153/2949 (5.2%) | 1.48 (0.79-2.56), p=0.17 | 1.72 (0.98-3.01), p=0.06 |  |
| Cardiovascular-related hospitalisation |  |  |  |  |
| HF in sinus rhythm | 557/1703 (32.7%) vs 2962/11865 (25.0%) | 1.29 (1.18-1.42), p<0.001 | 1.28 (1.16-1.42), p<0.001 | 0.14 |
| HF plus AF | 79/196 (40.3%) vs 879/2949 (29.8%) | 1.61 (1.26-2.03), p<0.001 | 1.56 (1.23-1.98), p<0.001 |  |
| Heart failure-related hospitalisation |  |  |  |  |
| HF in sinus rhythm | 470/1703 (27.6%) vs 1736/11865 (14.6%) | 1.94 (1.75-2.15), p<0.001 | 1.49 (1.33-1.66), p<0.001 | 0.07 |
| HF plus AF | 69/196 (35.2%) vs 606/2949 (20.5%) | 2.07 (1.59-2.66), p<0.001 | 2.05 (1.59-2.65), p<0.001 |  |

HF = heart failure; AF = atrial fibrillation; CI=confidence interval

**Figure 1:**
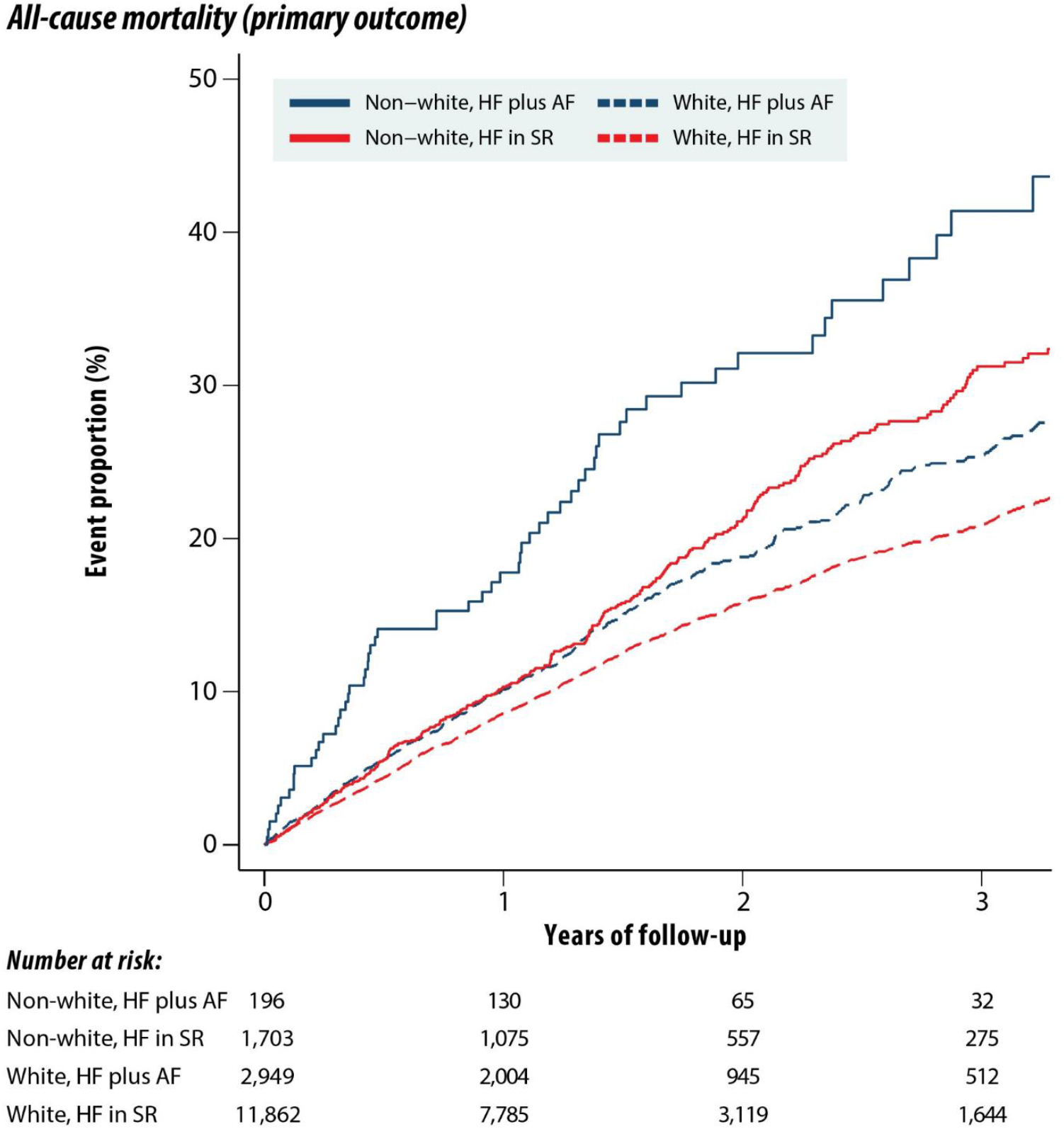
Kaplan-Meier curves for all-cause mortality stratified by ethnicity. Comparing trial participants with heart failure in sinus rhythm and heart failure plus AF. AF=atrial fibrillation; HF=heart failure; SR=sinus rhythm.

### Secondary outcomes

Crude event rates and adjusted HRs for each secondary outcome are presented in Table 2 and Figure 2, with consistent effects seen in the propensity-score matched analyses (Supplementary Table S5).

**Figure 2:**
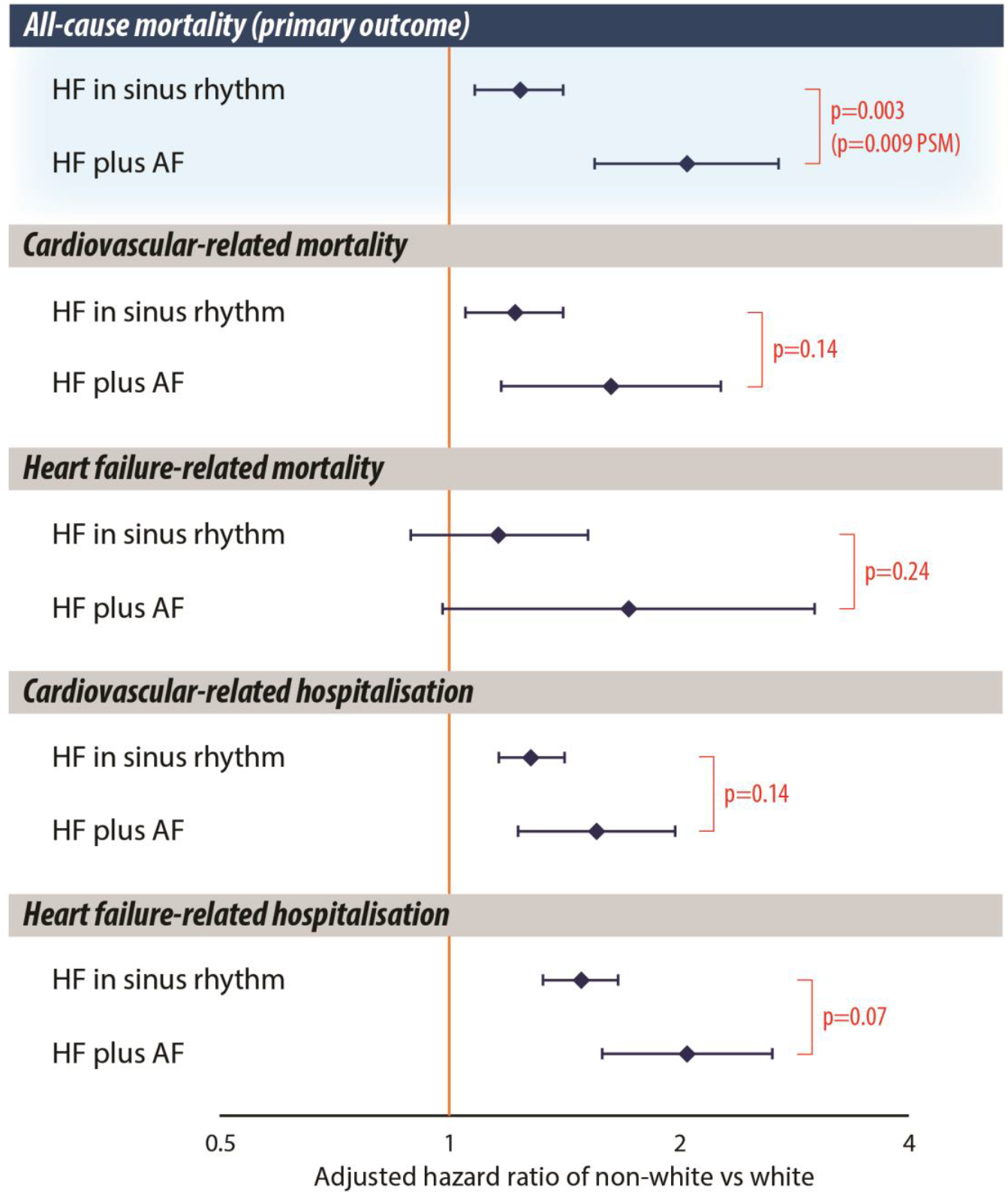
Forest plot of mortality and hospitalisation outcomes in white and non-white patients according to AF status. Adjusted hazard ratios with 95% confidence intervals and interaction p-values. Cox-proportional hazards models were adjusted for age, gender, left ventricular ejection fraction, New York Heart Association class, previous myocardial infarction, diabetes, and hypertension. AF=atrial fibrillation; HF=heart failure; PSM=propensity-score matched analysis.

The adjusted non-white versus white HR for CV-related death was 1.22 for HF in sinus rhythm (95% CI 1.05-1.41; p=0.007) and 1.63 for HF plus AF (95% CI 1.17-2.27; p=0.004); interaction p=0.14. For HF-related death, the adjusted HR was 1.16 for HF in sinus rhythm (95% CI 0.89-1.52; p=0.28) and 1.72 for HF plus AF (95% CI 0.98-3.01; p=0.06), with no statistical divergence (p=0.24).

For CV-related hospitalisation, the adjusted non-white versus white HR was 1.28 for HF in sinus rhythm (95% CI 1.16-1.42; p<0.001) and 1.56 for HF plus AF (95% CI 1.23-1.98; p<0.001); interaction p=0.14. The adjusted HR for HF-related hospitalisation was 1.49 for HF in sinus rhythm (95% CI 1.33-1.66; p<0.001) and 2.05 for HF plus AF (95% CI 1.59-2.65; p<0.001); interaction p=0.07.

### Exploratory analysis of therapeutic efficacy

Treatment effects were not significant for beta-blockers versus placebo in HF plus AF, or for spironolactone versus placebo in HF with sinus rhythm or AF (Supplementary Table S6). Analysis of beta-blockers for HF in sinus rhythm demonstrated significantly lower all-cause mortality versus placebo for white patients (HR 0.70, 95% CI 0.63-0.78; p<0.001), which was not seen in the non-white group (HR 1.00; 95% CI 0.79-1.27; p=0.99); p-interaction for ethnicity=0.008 and propensity-matched interaction p=0.001 (Figure 3 and Supplementary Table S7). The significant differential impact of beta-blockers versus placebo according to ethnicity was also seen for CV-related and HF-related mortality (Figure 3 and Supplementary Table S7).

**Figure 3:**
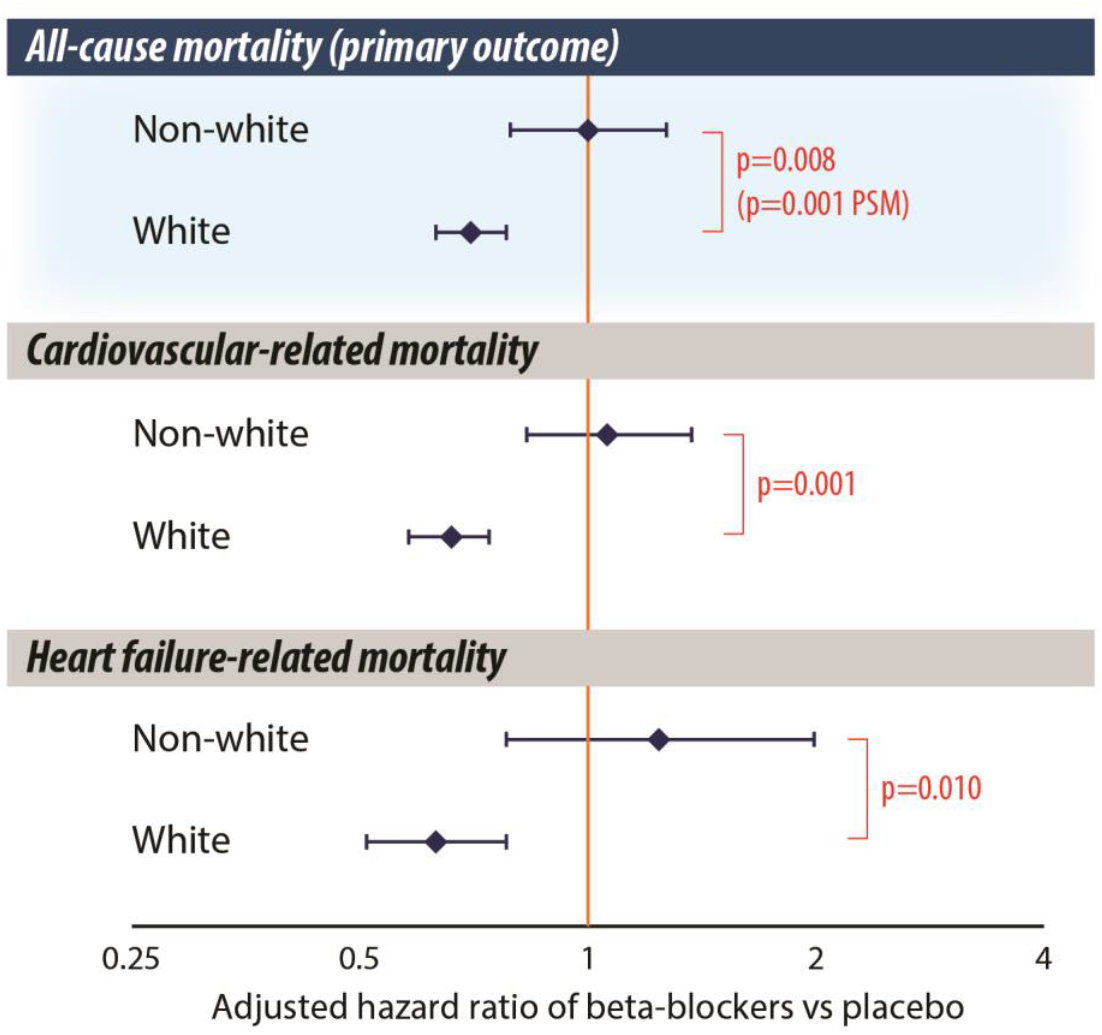
Exploratory analysis of mortality in white and non-white patients according to beta-blocker use in sinus rhythm. Adjusted hazard ratios with 95% confidence intervals and interaction p-values. Cox-proportional hazards models adjusted for age, gender, left ventricular ejection fraction, New York Heart Association class, previous myocardial infarction, diabetes, and hypertension. PSM=propensity-score matched analysis.

## Discussion

Using individual patient data from RCTs, this study found that the combination of HF and AF worsens the disparity in all-cause mortality seen between non-white and white patients. There was a quarter increase in hazard for non-white patients with HF alone, versus a 2-fold increase in hazard compared to white patients for those with HF plus AF. The incidence of the secondary outcomes CV-related death, HF-related death, CV-related hospitalisation and HF-related hospitalisation were also significantly higher in non-white patients.

Ethnicity plays a major and complex role in healthcare, impacting on presentation, diagnosis, treatment and outcomes. It is also closely related to a range of confounding factors that interrupt any direct association with reported cardiovascular events and death. For example, individuals in different ethnic groups can present later with their initial symptoms, may be less likely to seek healthcare assistance, or face barriers to access healthcare.^25^ Previous observational studies have suggested that non-white patients are significantly less likely to be discharged on appropriate medical therapy,^8^ or receive interventional procedures such as cardiac resynchronization therapy for HF and catheter ablation for AF.^9, 26^ All of these factors (and more) will contribute to the worse prognosis seen in non-white patients. The concept that a combination of health conditions could exacerbate health inequality has been demonstrated beyond cardiology. In both the United States and United Kingdom, studies have shown the mortality rate associated with multimorbidity is higher in certain ethnic groups compared to white patients.^27^

To counter some of the biases of observational data, this study only included randomised trials, with double-blinding, intention-to-treat analysis and independent adjudication of outcome events. Further, the use of individual patient-level data allowed for adjustment and matching with time-to-event analyses. However, none of the trials were randomised on the basis of ethnicity, so residual confounding for the aforementioned confounders will still apply and we clearly demonstrated a more severe HF phenotype at baseline in non-white participants. Propensity-matched analysis is not a surrogate for randomisation, and the more limited matched sample size may not be powered to detect interaction by ethnicity. The exploratory analysis of treatment effect by ethnicity was only possible for beta-blockers in patients with HF in sinus rhythm, as there was no significant benefit in the whole group for beta-blockers versus placebo in HF plus AF^2, 3^, or spironolactone versus placebo in any group.^11^ In direct contrast to the white participants, non-white patients appeared to lack benefit from beta-blockers, although this may have been related to later presentation as evidenced by lower LVEF, more symptoms and greater need for diuretics. We found no difference in randomisation to the intervention groups related to ethnicity, although there were differences in other medical therapy, notably less aldosterone antagonists in the non-white participants. Unlike prior observational studies, the non-white group in these RCTs had higher levels of ACEi/ARB and oral anticoagulation prescription. An additional factor in treatment response that we could not assess were genetic polymorphisms that can affect beta-blocker response^28^, and have different prevalence across ethnic groups.^29^ There are also numerous differences related to ethnicity in the pharmacokinetic and pharmacodynamic response to many other cardiovascular therapies.^30^

This study was limited by the numbers of non-white participants in these trials, requiring a combined assessment of non-white versus white, as opposed to individual ethnicities. By amalgamating these groups into a single cohort, any variation in outcome rates between different non-white ethnicities are obscured. Socioeconomic status, and related factors such as nutrition and salt intake, are other important confounders with ethnicity, but information on these variables was not collected in the component trials. Further, the trials incorporated are now somewhat dated, with additional medical therapy such as cardiac resynchronisation therapy, neprilysin inhibition and sodium-glucose cotransporter-2 inhibitors not available when these trials were conducted. However, all of the latter therapies are based on the foundation of ACEi, beta-blockers and aldosterone antagonists which were the subject of the included trials. Extrapolation of these findings to recent therapies and real-world data sources is a suggested next step.

The results of this study should spur action to improve research and the management of HF in relation to ethnic disparity. Concerted effort is needed to address the under-representation of ethnic minorities in medical research. A broader awareness of prescription and therapeutic biases in routine practice is critical. Deeper focus is required by all healthcare practitioners of the individual issues that each patient may face in order to achieve optimal outcomes, reduce mortality and morbidity, and reduce the substantial healthcare and social costs of heart failure.

## Conclusion

Analysis of patient-level data from double-blind randomised trials demonstrated a substantial excess of mortality for non-white patients with HF, compared to those of white ethnicity. The divergence in clinical outcomes was exacerbated in patients with additional AF, which is one of the most common concomitant conditions in HF. Further research is warranted to understand if different non-white groups have varying levels of risk and treatment efficacy.

## Data Availability

All data produced in the present study are available upon reasonable request to the corresponding author.

## Acknowledgements

We are indebted to the patients of the included trials, the steering committees, and other members of the Beta-blockers in Heart Failure Collaborative Group: Milton Packer MD (USA), Jane Holmes PhD (UK), Andrew JS Coats MD DSc (UK), John Wikstrand MD PhD (Sweden), Bert Andersson MD PhD (Sweden), Dirk J van Veldhuisen MD (the Netherlands), Thomas G von Lueder MD PhD (Norway), Frank Ruschitzka MD (Switzerland), John G Cleland MD (UK), Alan S Rigby MSc (UK), Marcelo C Shibata MD (Canada), Peter D Collins MD (UK), John Kjekshus PhD (Norway), Hans Wedel PhD (Sweden), John JV McMurray MD (UK), Stefan D. Anker MD PhD (Germany), and Luis Manzano MD (Spain). This work is dedicated to the memory of Douglas Altman (1948-2018; University of Oxford, UK), Philip Poole-Wilson (1943-2009; Imperial College London, UK) and Henry Krum (1958-2015; Monash University, Melbourne, Australia). BB-meta-HF was only possible with full access to trial data and support provided by AstraZeneca, GlaxoSmithKline, Menarini Farmaceutica and Merck Serono; with thanks. This paper was prepared using the Beta-Blocker Evaluation Survival (BEST) and Treatment of Preserved Cardiac Function Heart Failure with an Aldosterone Antagonist (TOPCAT) research materials obtained from the National Heart, Lung, and Blood Institute (NHLBI) Biologic Specimen and Data Repository Information Coordinating Center (BioLINCC). The paper does not reflect the opinions or views of the BEST or TOPCAT investigators, or the NHLBI. The card*AI*c group (Application of Artificial Intelligence to Routine Healthcare Data to Benefit Patients with Cardiovascular Disease) includes researchers at the University of Birmingham and University Hospitals Birmingham NHS Foundation Trust: Animesh Acharjee, David Brind, Karina V Bunting, Jun Yu Chen, Jinming Duan, Georgios V. Gkoutos, Xin Guan, Andreas Karwath, Arcella Lim, Alastair Mobley, Daniella Okyere, Alex Thorley, Maximina Ventura, Xiaoxia Wang and Yuting Zhang.

## Graphical Abstract

Analysis of individual patient data from heart failure trials indicates an ethnic disparity in clinical outcomes, which is exacerbated by cardiovascular multimorbidity.

AF=atrial fibrillation; HF=heart failure; HR=hazard ratio; SR=sinus rhythm.

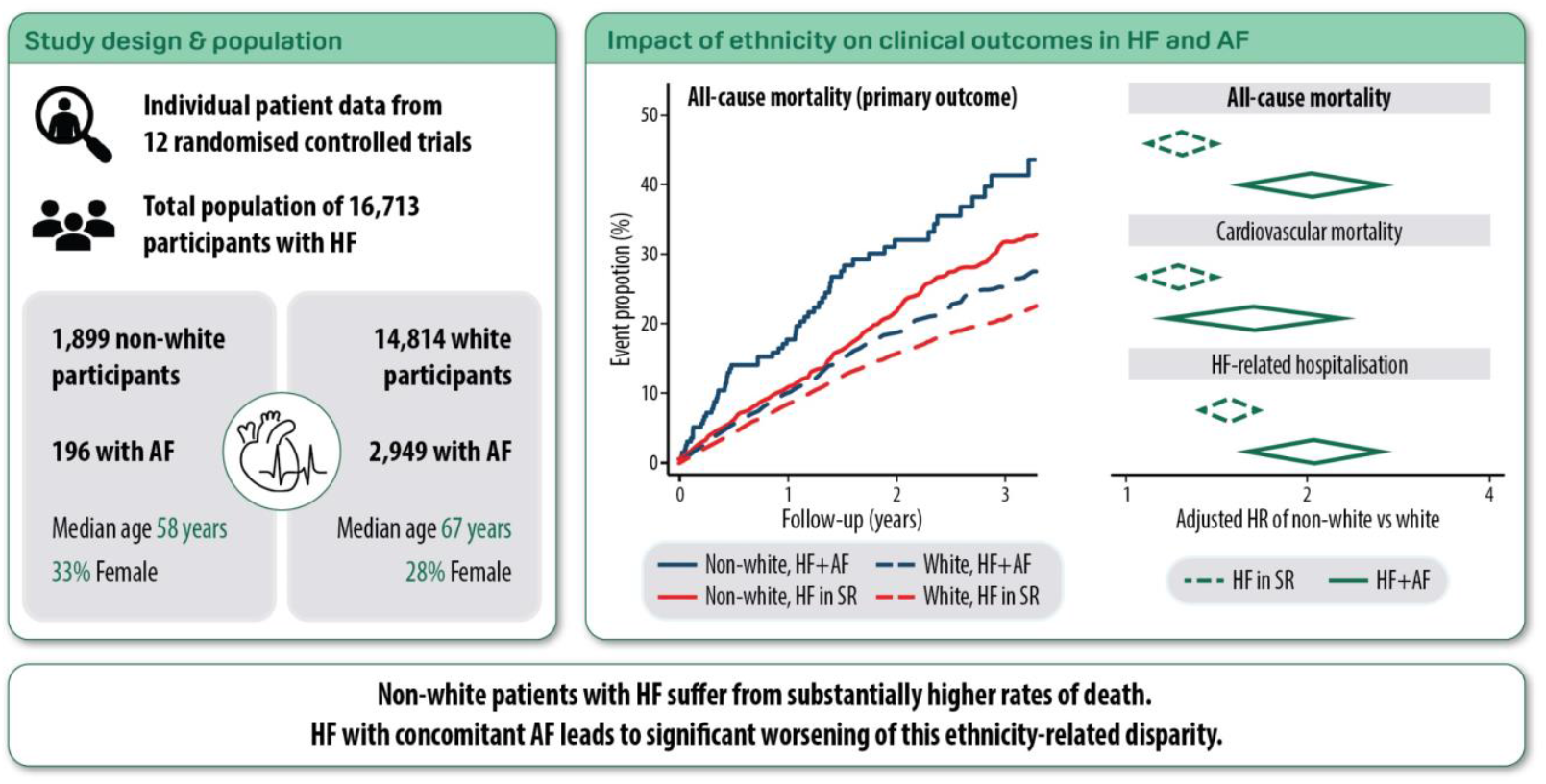

## References

1. Ţica O, Khamboo W, Kotecha D. Breaking the Cycle of Heart Failure With Preserved Ejection Fraction and Atrial Fibrillation. Card Fail Rev. 2022;8:e32. 10.15420/cfr.2022.03.

2. Kotecha D, Holmes J, Krum H, Altman DG, Manzano L, Cleland JGF, et al. Efficacy of β blockers in patients with heart failure plus atrial fibrillation: An individual-patient data meta-analysis. The Lancet. 2014;384:2235–2243. 10.1016/S0140-6736(14)61373-8.

3. Karwath A, Bunting KV, Gill SK, Tica O, Pendleton S, Aziz F, et al. Redefining beta-blocker response in heart failure patients with sinus rhythm and atrial fibrillation: a machine learning cluster analysis. Lancet. 2021;398:1427–1435. 10.1016/S0140-6736(21)01638-X.

4. Hayanga B, Stafford M, Bécares L. Ethnic inequalities in multiple long-term health conditions in the United Kingdom: a systematic review and narrative synthesis. BMC Public Health. 2023;23:178. 10.1186/s12889-022-14940-w.

5. Banerjee A. Ethnicity, heart failure and the prevention continuum: time to act. Heart. 2020;106:631–633. 10.1136/heartjnl-2019-316293.

6. Magnani JW, Norby FL, Agarwal SK, Soliman EZ, Chen LY, Loehr LR, et al. Racial Differences in Atrial Fibrillation-Related Cardiovascular Disease and Mortality: The Atherosclerosis Risk in Communities (ARIC) Study. JAMA Cardiol. 2016;1:433–41. 10.1001/jamacardio.2016.1025.

7. Virani SS, Alonso A, Benjamin EJ, Bittencourt MS, Callaway CW, Carson AP, et al. Heart Disease and Stroke Statistics-2020 Update: A Report From the American Heart Association. Circulation. 2020;141:e139–e596. 10.1161/cir.0000000000000757.

8. Thomas KL, Piccini JP, Liang L, Fonarow GC, Yancy CW, Peterson ED, et al. Racial differences in the prevalence and outcomes of atrial fibrillation among patients hospitalized with heart failure. J Am Heart Assoc. 2013;2:e000200. 10.1161/jaha.113.000200.

9. Bhatia S, Qazi M, Erande A, Shah K, Amin A, Patel P, et al. Racial Differences in the Prevalence and Outcomes of Atrial Fibrillation in Patients Hospitalized With Heart Failure. Am J Cardiol. 2016;117:1468–73. 10.1016/j.amjcard.2016.02.016.

10. Kotecha D, Manzano L, Altman DG, Krum H, Erdem G, Williams N, et al. Individual patient data meta-analysis of beta-blockers in heart failure: rationale and design. Syst Rev. 2013;2:7. 10.1186/2046-4053-2-7.

11. Pitt B, Pfeffer MA, Assmann SF, Boineau R, Anand IS, Claggett B, et al. Spironolactone for heart failure with preserved ejection fraction. N Engl J Med. 2014;370:1383–92. 10.1056/NEJMoa1313731.

12. Eichhorn EJ, Domanski MJ, Krause-Steinrauf H, Bristow MR, Lavori PW. A trial of the beta-blocker bucindolol in patients with advanced chronic heart failure. N Engl J Med. 2001;344:1659–67. 10.1056/nejm200105313442202.

13. Kotecha D, Manzano L, Krum H, Rosano G, Holmes J, Altman DG, et al. Effect of age and sex on efficacy and tolerability of β blockers in patients with heart failure with reduced ejection fraction: individual patient data meta-analysis. Bmj. 2016;353:i1855. 10.1136/bmj.i1855.

14. Leuven E, Sianesi B. PSMATCH2: Stata module to perform full Mahalanobis and propensity score matching, common support graphing, and covariate imbalance testing. Statistical Software Components 2018

15. A randomized trial of beta-blockade in heart failure. The Cardiac Insufficiency Bisoprolol Study (CIBIS). CIBIS Investigators and Committees. Circulation. 1994;90:1765–73. 10.1161/01.cir.90.4.1765.

16. Cleland JG, Pennell DJ, Ray SG, Coats AJ, Macfarlane PW, Murray GD, et al. Myocardial viability as a determinant of the ejection fraction response to carvedilol in patients with heart failure (CHRISTMAS trial): randomised controlled trial. Lancet. 2003;362:14–21. https://doi.org/S0140673603138019 [pii].

17. Dargie HJ. Effect of carvedilol on outcome after myocardial infarction in patients with left-ventricular dysfunction: the CAPRICORN randomised trial. Lancet. 2001;357:1385–90. 10.1016/s0140-6736(00)04560-8.

18. Flather MD, Shibata MC, Coats AJ, Van Veldhuisen DJ, Parkhomenko A, Borbola J, et al. Randomized trial to determine the effect of nebivolol on mortality and cardiovascular hospital admission in elderly patients with heart failure (SENIORS). Eur Heart J. 2005;26:215–25.

19. Packer M, Bristow MR, Cohn JN, Colucci WS, Fowler MB, Gilbert EM, et al. The effect of carvedilol on morbidity and mortality in patients with chronic heart failure. U.S. Carvedilol Heart Failure Study Group. N Engl J Med. 1996;334:1349–55.

20. Packer M, Coats AJ, Fowler MB, Katus HA, Krum H, Mohacsi P, et al. Effect of carvedilol on survival in severe chronic heart failure. N Engl J Med. 2001;344:1651–8.

21. Waagstein F, Bristow MR, Swedberg K, Camerini F, Fowler MB, Silver MA, et al. Beneficial effects of metoprolol in idiopathic dilated cardiomyopathy. Metoprolol in Dilated Cardiomyopathy (MDC) Trial Study Group. Lancet. 1993;342:1441–6.

22. The Cardiac Insufficiency Bisoprolol Study II (CIBIS-II): a randomised trial. Lancet. 1999;353:9–13. https://doi.org/S0140673698111819 [pii].

23. Effect of metoprolol CR/XL in chronic heart failure: Metoprolol CR/XL Randomised Intervention Trial in Congestive Heart Failure (MERIT-HF). Lancet. 1999;353:2001–7. https://doi.org/S0140673699044402 [pii].

24. Randomised, placebo-controlled trial of carvedilol in patients with congestive heart failure due to ischaemic heart disease. Australia/New Zealand Heart Failure Research Collaborative Group. Lancet. 1997;349:375–80. 10.1016/S0140-6736(97)80008-6.

25. Williams DR, Rucker TD. Understanding and addressing racial disparities in health care. Health Care Financ Rev. 2000;21:75–90.

26. Sridhar AR, Yarlagadda V, Parasa S, Reddy YM, Patel D, Lakkireddy D, et al. Cardiac Resynchronization Therapy: US Trends and Disparities in Utilization and Outcomes. Circ Arrhythm Electrophysiol. 2016;9:e003108. 10.1161/circep.115.003108.

27. Quiñones AR, Newsom JT, Elman MR, Markwardt S, Nagel CL, Dorr DA, et al. Racial and Ethnic Differences in Multimorbidity Changes Over Time. Med Care. 2021;59:402–409. 10.1097/mlr.0000000000001527.

28. Bristow MR, Murphy GA, Krause-Steinrauf H, Anderson JL, Carlquist JF, Thaneemit-Chen S, et al. An alpha2C-adrenergic receptor polymorphism alters the norepinephrine-lowering effects and therapeutic response of the beta-blocker bucindolol in chronic heart failure. Circ Heart Fail. 2010;3:21–8. 10.1161/circheartfailure.109.885962.

29. Taylor MR, Sun AY, Davis G, Fiuzat M, Liggett SB, Bristow MR. Race, common genetic variation, and therapeutic response disparities in heart failure. JACC Heart Fail. 2014;2:561–72. 10.1016/j.jchf.2014.06.010.

30. Tamargo J, Kaski JC, Kimura T, Barton JC, Yamamoto K, Komiyama M, et al. Racial and ethnic differences in pharmacotherapy to prevent coronary artery disease and thrombotic events. Eur Heart J Cardiovasc Pharmacother. 2022;8:738–751. 10.1093/ehjcvp/pvac040.

