## Supplementary for "Combination of heart failure and atrial fibrillation worsens ethnicity-related disparity: An individual patient-level meta-analysis of randomised trials"

Online Supplement

### Supplementary Table S1: Breakdown of population by ethnicity and rhythm status

| **Ethnicity** | **HF in sinus rhythm** | **HF plus AF** | **Total** |
| --- | --- | --- | --- |
| White/Caucasian | 11,865 (87%) | 2,949 (94%) | 14,814 (89%) |
| African/African-American | 1,226 (9%) | 141 (4%) | 1,367 (8%) |
| Asian* | 144 (1%) | 20 (1%) | 164 (1%) |
| Other** | 333 (2%) | 35 (1%) | 368 (2%) |
| **Total** | **13,568** | **3,145** | **16,713** |

AF=atrial fibrillation; HF=heart failure

*Asian ethnicity refers to patients from South Asia, Southeast Asia, and East Asia

**An ethnicity of ‘Other’ was used for patients whose ethnic background was not represented by the other 3 categories, including: Hispanic, Middle Eastern, Native American, Indigenous, and people of the Pacific Islands

### **Supplementary Table S2: Baseline characteristics for HF in sinus rhythm**

| **Baseline characteristic** | **Non-white**  **(n=1,703)** | **White**  **(n=11,865)** | **p-value** |
| --- | --- | --- | --- |
| Intervention arm, n (%) | 884 (51.9%) | 6,048 (51.0%) | 0.47 |
| Women, n (%) | 582 (34.2%) | 3,348 (28.2%) | <0.001 |
| Age, median years (IQR) | 57.1 (48.4-66.0) | 65.9 (57.0-73.0) | <0.001 |
| BMI, median kg/m^2^ (IQR) | 28.0 (24.2-32.9) | 27.0 (24.3-30.4) | <0.001 |
| Systolic BP, median mmHg (IQR) | 120 (108-135) | 125 (112-139) | <0.001 |
| Diastolic BP, median mmHg (IQR) | 75 (68-82) | 76.7 (70-81.7) | 0.011 |
| Heart rate, mean (SD) | 82.5 (13.6) | 78.6 (12.4) | <0.001 |
| LVEF, median (IQR) | 0.24 (0.18-0.33) | 0.30 (0.22-0.37) | <0.001 |
| NYHA Class III/IV, n (%) | 1,216 (71.8%) | 6,167 (52.0%) | <0.001 |
| Comorbidities |  | | |
| Previous myocardial infarction, n (%) | 608 (35.7%) | 6,858 (57.8%) | <0.001 |
| Previous stroke, n (%) | 63 (6.9%) | 516 (6.1%) | 0.35 |
| Previous CABG or PCI, n (%) | 313 (18.5%) | 2,721 (23.6%) | <0.001 |
| Hypertension, n (%) | 1,167 (69.9%) | 6,017 (52.2%) | <0.001 |
| Peripheral arterial disease, n (%) | 140 (8.6%) | 981 (8.7%) | 0.89 |
| Diabetes mellitus, n (%) | 681 (40.8%) | 3,017 (26.0%) | <0.001 |
| Medication |  |  |  |
| Digoxin, n (%) | 1,201 (72.0%) | 5,074 (44.1%) | <0.001 |
| ACE inhibitor or ARB, n (%) | 1,413 (95.9%) | 9,688 (94.5%) | 0.020 |
| Calcium channel blocker, n (%) | 167 (10.1%) | 1,278 (11.0%) | 0.24 |
| Aldosterone antagonist, n (%) | 823 (7.1%) | 76 (4.6%) | <0.001 |
| Oral anticoagulant, n (%) | 513 (30.7%) | 2,773 (24.1%) | <0.001 |
| Diuretic, n (%) | 1,373 (93.2%) | 8,264 (80.6%) | <0.001 |
| Statin, n (%) | 407 (25.0%) | 3043 (27.0%) | 0.09 |

ACE = angiotensin-converting enzyme; ARB = angiotensin receptor blocker. BMI = body mass index; BP = blood pressure; CABG = coronary artery bypass grafting; IQR = interquartile range; LVEF = left ventricular ejection fraction; NYHA = New York Heart Failure Association; PCI = percutaneous coronary intervention; SD = standard deviation;

Missing data report: BMI n=19 non-white, n=104 white; systolic BP n=8 non-white, n=59 white; diastolic BP n=11 non-white, n=61 white; heart rate n=1 non-white, n=12 white; LVEF n=12 non-white, n=49 white; NYHA n=9 non-white, n=63 white; MI n=0 non-white, n=1 white; stroke n=788 non-white, n=3402 white; CABG/PCI n=14 non-white, n=343 white; hypertension n=34 non-white, n=349 white; peripheral arterial disease n=76 non-white, n=606 white; diabetes mellitus n=34 non-white, n=259 white; digoxin n=34 non-white, n=349 white; ACE inhibitor or ARB n=230 non-white, n=1609 white; calcium channel blocker n=42 non-white, n=261 white; aldosterone antagonist n=34 non-white, n=349 white; oral anticoagulant n=34 non-white, n=350 white, diuretic n=58 non-white, n=644 white; statin n=4 non-white, n=60 white.

### **Supplementary Table S3: Baseline characteristics for HF plus AF**

| **Baseline characteristic** | **Non-White**  **(n=196)** | **White**  **(n=2,949)** | **P-value** |
| --- | --- | --- | --- |
| Intervention arm, n (%) | 102 (52.0%) | 1,462 (49.6%) | 0.50 |
| Women, n (%) | 52 (26.5%) | 741 (25.1%) | 0.66 |
| Age, median years (IQR) | 65.2 (57.0-74.0) | 71.0 (63.0-76.0) | <0.001 |
| BMI, median kg/m^2^ (IQR) | 27.8 (23.4-32.4) | 27.6 (24.7-31.2) | 0.86 |
| Systolic BP, median mmHg (IQR) | 119 (107.5-131.5) | 127 (115-140) | <0.001 |
| Diastolic BP, median mmHg (IQR) | 74.5 (65-80.8) | 78.3 (70-83) | 0.003 |
| Heart rate, mean (SD) | 77.1 (14.8) | 80.9 (14.1) | <0.001 |
| LVEF, median (IQR) | 0.30 (0.21-0.50) | 0.31(0.23-0.45) | 0.65 |
| NYHA Class III/IV, n (%) | 146 (74.9%) | 1,872 (63.7%) | 0.002 |
| Comorbidities |  | | |
| Previous myocardial infarction, n (%) | 54 (27.6%) | 1,047 (35.5%) | 0.023 |
| Previous stroke, n (%) | 10 (8.7%) | 177 (8.8%) | 0.98 |
| Previous CABG or PCI, n (%) | 29 (14.8%) | 481 (16.6%) | 0.51 |
| Hypertension, n (%) | 147 (75.0%) | 1,680 (57.0%) | <0.001 |
| Peripheral arterial disease, n (%) | 16 (8.3%) | 199 (6.9%) | 0.45 |
| Diabetes mellitus, n (%) | 66 (34.4%) | 738 (25.5%) | 0.007 |
| Medication |  |  |  |
| Digoxin, n (%) | 145 (74.0%) | 2,147 (72.8%) | 0.72 |
| ACE inhibitor or ARB, n (%) | 130 (94.2%) | 2,186 (94.8%) | 0.74 |
| Calcium channel blocker, n (%) | 35 (18.2%) | 352 (12.2%) | 0.014 |
| Aldosterone antagonist, n (%) | 11 (5.6%) | 420 (14.2%) | 0.001 |
| Oral anticoagulant, n (%) | 147 (75.0%) | 1,819 (61.7%) | <0.001 |
| Diuretic, n (%) | 136 (98.6%) | 2,395 (92.1%) | 0.005 |
| Statin, n (%) | 44 (22.9%) | 582 (20.1%) | 0.36 |

ACE = angiotensin-converting enzyme; ARB = angiotensin receptor blocker. BMI = body mass index; BP = blood pressure; CABG = coronary artery bypass grafting; IQR = interquartile range; LVEF = left ventricular ejection fraction; NYHA = New York Heart Failure Association; PCI = percutaneous coronary intervention; SD = standard deviation;

Missing data report: BMI n=4 non-white, n=18 white; systolic BP n=0 non-white, n=1 white; diastolic BP n=0 non-white, n=2 white; heart rate n=0 non-white, n=1 white; LVEF n=1 non-white, n=15 white; NYHA n=1 non-white, n=9 white; MI n=0 non-white, n=2 white; stroke n=81 non-white, n=928 white; CABG/PCI n=0 non-white, n=51 white; peripheral arterial disease n=4 non-white, n=60 white; diabetes mellitus n=4 non-white, n=60 white; ACE inhibitor or ARB n=58 non-white, n=644 white; calcium channel blocker n=4 non-white, n=60 white; diuretic n=58 non-white, n=644 white; statin n=4 non-white, n=60 white.

### **Supplementary Table S4: Baseline characteristics according to ethnicity after propensity score matching**

| **Variable** | **Non-white**  **(n=1354)** | **White**  **(n=1011)** | **p-value** |
| --- | --- | --- | --- |
| Age, mean years (SD) | 55.6 (12.2) | 55.4 (12.2) | 0.74 |
| Female gender, wt % | 30.7 | 30.6 | 0.97 |
| LVEF, mean % (SD) | 23.0 (7.7) | 23.2 (7.8) | 0.47 |
| NYHA class III/IV, wt % | 80.6 | 79.8 | 0.63 |
| BMI, mean kg/m^2^ (SD) | 28.3 (6.2) | 28.3 (5.5) | 0.95 |
| Heart rate, mean beats per min (SD) | 84.5 (12.8) | 84.6 (12.4) | 0.77 |
| Previous myocardial infarction, wt % | 37.6 | 36.6 | 0.58 |
| Diabetes, wt % | 38.5 | 41.0 | 0.18 |
| History of hypertension, wt % | 68.1 | 68.1 | 1.00 |
| Previous CABG or PCI, wt % | 17.7 | 19.0 | 0.37 |
| Digoxin, wt % | 84.3 | 83.5 | 0.60 |
| ACE inhibitor or ARB, wt % | 96.5 | 96.8 | 0.67 |
| Aldosterone antagonists, wt % | 5.2 | 5.4 | 0.86 |
| Anticoagulants, wt % | 33.1 | 30.3 | 0.12 |
| Diuretics, wt % | 94.2 | 94.2 | 1.00 |

ACE = angiotensin-converting enzyme; ARB = angiotensin receptor blocker; BMI = body mass index; CABG = coronary artery bypass grafting; LVEF = left ventricular ejection fraction; NYHA = New York Heart Failure Association; PCI = percutaneous coronary intervention; SD = standard deviation.

Wt % = weighted percentage

### Supplementary Table S5: Propensity score-matched analysis for primary and secondary outcomes

| **Outcome** | **Crude event numbers (%)** | **Propensity score matching ATT for non-white vs white (95% CI), p-value** | **P-value for comparison of sinus rhythm vs AF** |
| --- | --- | --- | --- |
| **All-cause mortality** | | | |
| HF in sinus rhythm | 330/1703 (19.4%) vs 1610/11865 (13.6%) | 0.024 (-0.009-0.057), p=0.17 | <0.001 |
| HF plus AF | 64/196 (32.7%) vs 532/2949 (18.0%) | 0.178 (0.07-0.286), p=0.001 |  |
| **Cardiovascular-related mortality** | | | |
| HF in Sinus rhythm | 278/1703 (16.3%) vs 1317/11865 (11.1%) | 0.016 (-0.015-0.047), p=0.33 | 0.021 |
| HF plus AF | 45/196 (23.0%) vs 432/2949 (14.6%) | 0.116 (0.012-0.022), p=0.028 |  |
| **Heart failure-related mortality** | | | |
| HF in Sinus rhythm | 75/1703 (4.4%) vs 376/11865 (3.2%) | ^-^0.007 (^-^0.027-0.013), p=0.46 | 0.16 |
| HF plus AF | 15/196 (7.7%) vs 153/2949 (5.2%) | 0.031 (^-^0.04-0.102), p=0.39 |  |
| **Cardiovascular-related hospitalisation** | | | |
| HF in Sinus rhythm | 557/1703 (32.7%) vs 2962/11865 (25.0%) | 0.039 (0.00-0.078), p=0.05 | 0.24 |
| HF plus AF | 79/196 (40.3%) vs 879/2949 (29.8%) | 0.101 (^-^0.024-0.226), p=0.12 |  |
| **Heart failure-related hospitalisation** | | | |
| HF in Sinus rhythm | 470/1703 (27.6%) vs 1736/11865 (14.6%) | 0.047 (0.010-0.084), p=0.014 | 0.031 |
| HF plus AF | 69/196 (35.2%) vs 606/2949 (20.5%) | 0.155 (0.035-0.275), p=0.011 |  |

AF=Atrial Fibrillation; ATT=Average Treatment Effect on the Treated; CI=Confidence Interval

### **Supplementary Table S6: Heart failure t**reatment **efficacy according to cardiac rhythm status.**

|  | **All-cause mortality adjusted hazard ratio (95% CI), p-value** | |
| --- | --- | --- |
|  | **Beta blockers vs Placebo** | **Spironolactone vs Placebo** |
| HF in sinus rhythm | 0.75 (0.67-0.82), p<0.001 | 0.90 (0.68-1.18), p=0.44 |
| HF plus AF | 0.82 (0.71-1.04), p=0.13 | 0.80 (0.55-1.16), p=0.24 |

CI=Confidence Interval

### Supplementary Table S7: Cox regression and propensity score-matched analysis of beta blocker efficacy according to ethnicity in patients with HF in sinus rhythm

| **Beta-blocker vs Placebo:** | **Crude event numbers (%)** | **Adjusted hazard ratio (95% CI), p-value** | **Interaction p-value for ethnicity** | **Propensity score matched ATT (95% CI),**  **p-value** | **p-value*** |
| --- | --- | --- | --- | --- | --- |
| **All-cause mortality** | | | | | |
| Non-White | 170/884 (19.2%) vs 160/819 (19.5%) | 1.00 (0.79-1.27), p=0.99 | 0.008 | 0.018 (^-^0.04-0.077), p=0.53 | 0.030 |
| White | 706/6048 (11.7%) vs 904/5817 (15.5%) | 0.70 (0.63-0.78), p<0.001 |  | 0.043 (^-^0.063-^-^0.024), p<0.001 |  |
| **Cardiovascular-related mortality** | | | | | |
| Non-White | 146/884 (16.5%) vs 132/819 (16.1%) | 1.06 (0.83-1.37), p=0.83 | 0.001 | 0.033 (-0.023-0.088), p=0.25 | 0.002 |
| White | 556/6048 (9.2%) vs 761/5817 (13.1%) | 0.66 (0.58-0.74), p<0.001 |  | 0.048 (^-^0.067-^-^0.03), p<0.001 |  |
| **Heart failure-related mortality** | | | | | |
| Non-White | 42/884 (4.8%) vs 33/819 (4.0%) | 1.24 (0.78-1.99), p=0.37 | 0.010 | 0.024 (^-^0.005-0.054), p=0.11 | 0.003 |
| White | 153/6048 (2.5%) vs 223/5817 (3.8%) | 0.63 (0.51-0.78), p<0.001 |  | 0.018 (^-^0.028-^-^0.007), p=0.001 |  |

ATT=Average Treatment Effect on the Treated; CI=Confidence Interval

*T-test comparing the ATT of Non-White patients with White patient

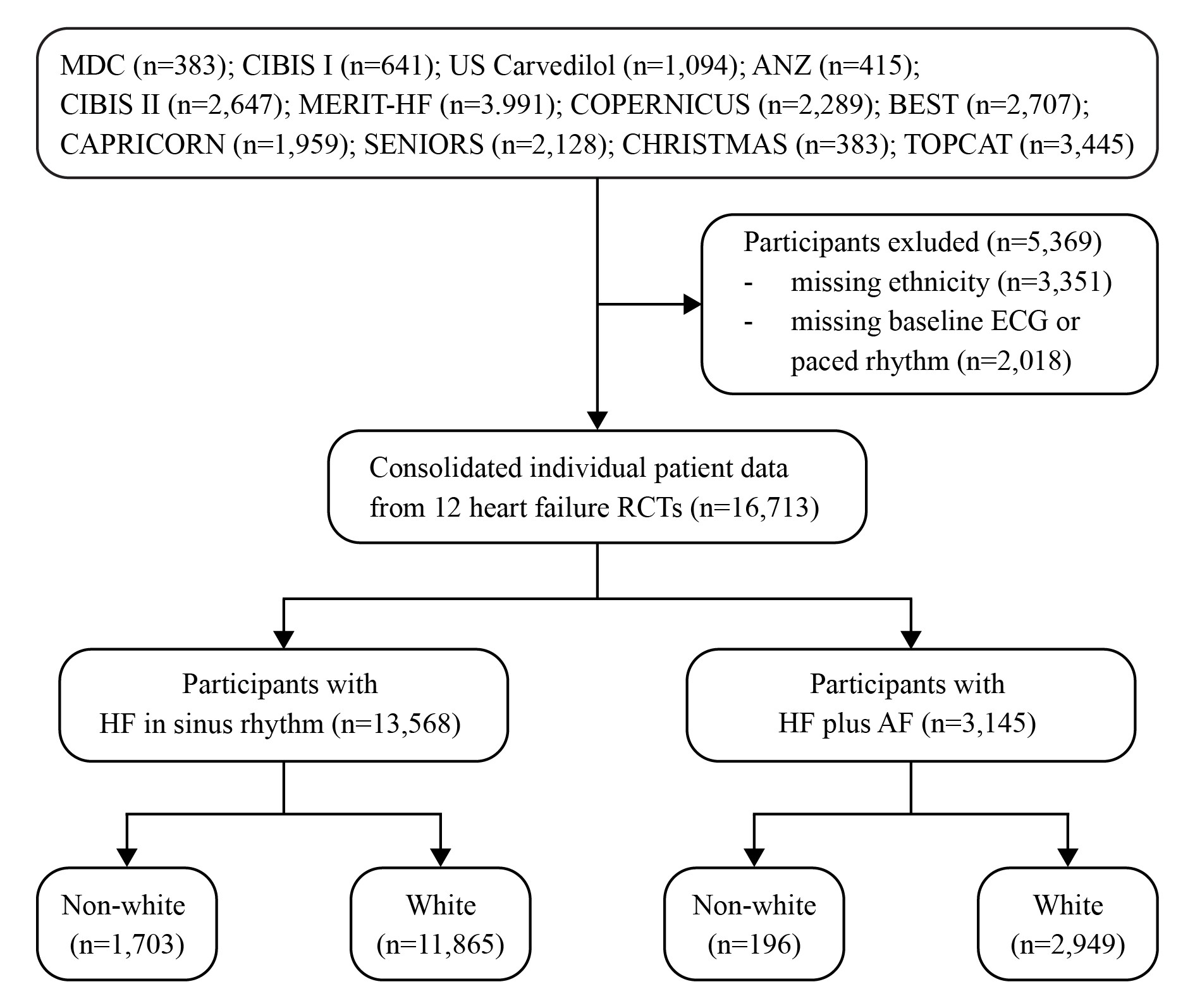

### Supplementary Figure S1: Participant flow diagram

AF=Atrial Fibrillation; ANZ=Australia/New Zealand Heart Failure Study; BEST=Beta-Blocker Evaluation Survival Trial; CAPRICORN=Carvedilol Post-Infarct Survival Control in Left Ventricular Dysfunction Study; CHRISTMAS=Carvedilol Hibernating Reversible Ischaemia Trial: Marker of Success Study; CIBIS I=Cardiac Insufficiency Bisoprolol Study; CIBIS II=Cardiac Insufficiency Bisoprolol Study II; COPERNICUS=Carvedilol Prospective Randomised Cumulative Survival Study; ECG=Electrocardiogram; HF=Heart Failure; MDC=Metoprolol in Dilated Cardiomyopathy Trial;

MERIT-HF=Metoprolol CR/XL Randomised Intervention Trial in Congestive Heart Failure; RCT=Randomised Controlled Trial; SENIORS=Study of the Effects of Nebivolol Intervention in Outcomes and Rehospitalisation in Seniors with Heart Failure Study; TOPCAT=Treatment of Preserved Cardiac Function Heart Failure with an Aldosterone Antagonist

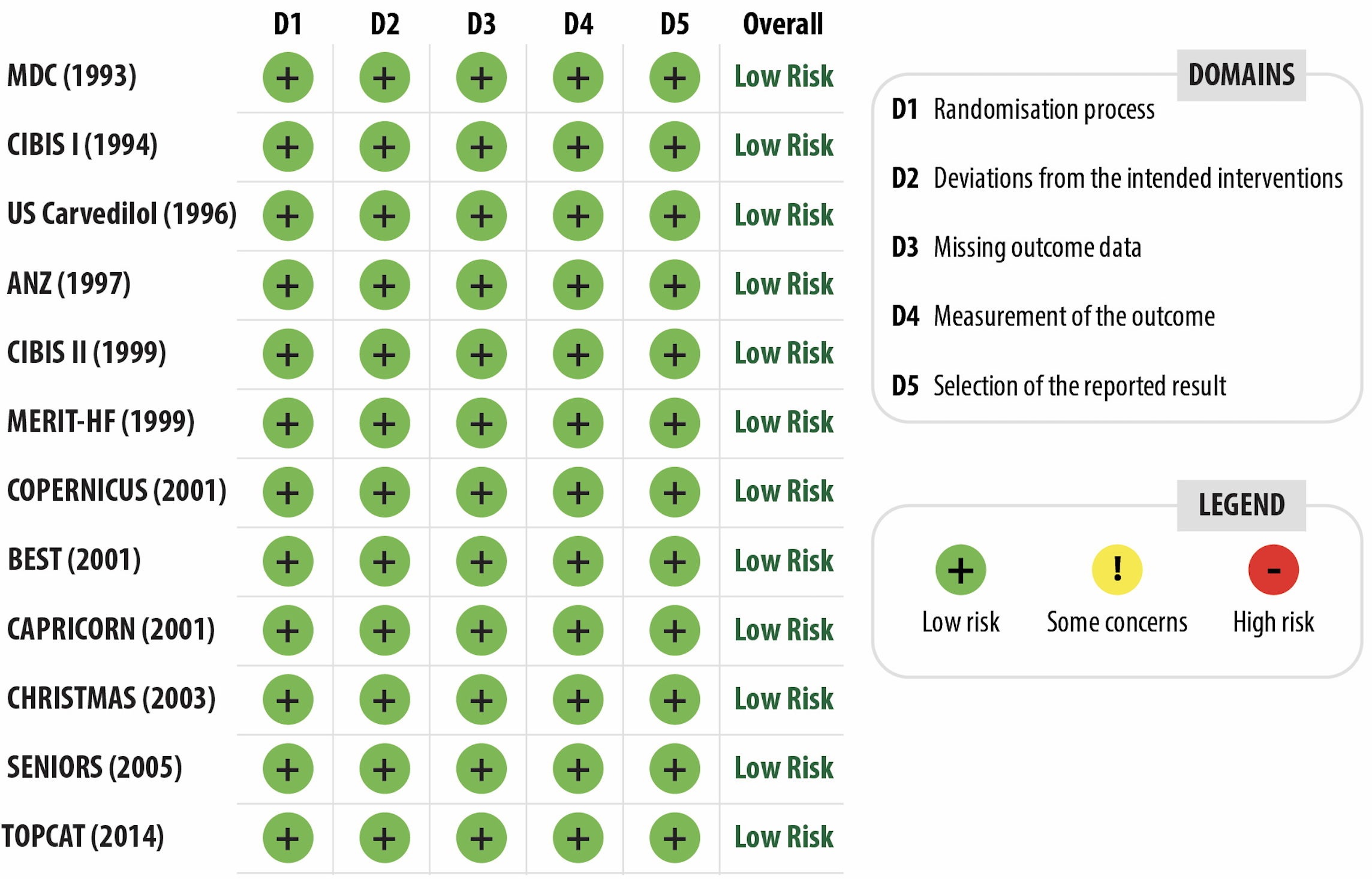

### Supplementary Figure S2: Risk of Bias Assessment

ANZ=Australia/New Zealand Heart Failure Study; BEST=Beta-Blocker Evaluation Survival Trial; CAPRICORN=Carvedilol Post-Infarct Survival Control in Left Ventricular Dysfunction Study; CHRISTMAS=Carvedilol Hibernating Reversible Ischaemia Trial: Marker of Success Study; CIBIS I=Cardiac Insufficiency Bisoprolol Study; CIBIS II=Cardiac Insufficiency Bisoprolol Study II;

COPERNICUS=Carvedilol Prospective Randomised Cumulative Survival Study; MDC=Metoprolol in Dilated Cardiomyopathy Trial; MERIT-HF=Metoprolol CR/XL Randomised Intervention Trial in Congestive Heart Failure; SENIORS=Study of the Effects of Nebivolol Intervention in Outcomes and Rehospitalisation in Seniors with Heart Failure Study; TOPCAT=Treatment of Preserved Cardiac Function Heart Failure with an Aldosterone Antagonist;
